# Impact of Hypertension on Progression and Prognosis in Patients with COVID-19 *A Retrospective Cohort Study in 1031 Hospitalized Cases in Wuhan, China*

**DOI:** 10.1101/2020.06.14.20125997

**Authors:** Hesong Zeng, Tianlu Zhang, Xingwei He, Yuxin Du, Yan Tong, Xueli Wang, Weizhong Zhang, Yin Shen

**Affiliations:** Department of Cardiology, Tongji Hospital, Tongji Medical College, Huazhong University of Science and Technology, Wuhan, China; Medical Research Institute, Renmin Hospital of Wuhan University, Wuhan University, Wuhan, China; Institute of Central China Development, Wuhan University, Wuhan, China; Shanghai Institute of Hypertension, Ruijin Hospital, Shanghai Jiaotong University, Shanghai, China

**Keywords:** COVID-19, SARS-COV-2, Comorbidities, Hypertension, CCB, in-hospital case-fatality rate

## Abstract

**Objectives:** The main aim of our study is to observe clinical characteristics and effects of antihypertensive drugs in different hospitalized populations, and to further provide evidence to explore causes and specific clinical markers of the aggravation of COVID-19 in patients with underlying hypertension.

**Design:** This was a retrospective cohort study focusing on the clinical data of COVID-19 inpatients admitted at the early stage of pandemic.

**Setting:** A single center study conducted in Tongji Hospital, Tongji Medical College of Huazhong university of Science and Technology (Wuhan, China).

**Participants:** All 1031 inpatients diagnosed with COVID-19 according to *Prevention and control Scheme for Novel Coronavirus Pneumonia* published by National Health Commission of the People’s Republic of China and WHO interim guidance in Tongji hospital (Wuhan, China), from January 27, 2020, to March 8, 2020 with the cutoff date being March 30, 2020, were included in this study.

**Main outcome measures:** Demographic data, medical history, clinical symptoms and signs, laboratory findings, chest computed tomography (CT), treatment, and clinical outcomes were extracted from electronic medical records.

**Results:** 1031 COVID-19 inpatients were included in this study, of whom 866 were discharged and 165 were deceased in hospital. 73% of 165 deceased patients had chronic comorbidities. Patients with underlying diseases showed CFR 2.8 times as that of patients without. Senility and males were observed to be main risk factors for increased in-hospital case-fatality rate, with the odds ratio in multivariable regression being 2.94 (95%CI: 2.00 to 4.33; P <0.001) and 2.47 (95%CI: 1.65 to 3.70; P <0.001), respectively. The odds ratio of cases with composite endpoints for patients with simple hypertension was 1.53 (95%CI: 1.07 to 2.17; P=0.019). Senile patients with hypertension were proved to be at high risk early in the disease, which might be associated with the level of CRP, LDH, and eGFR. The odds ratio of case-fatality rate for patients with hypertension taking CCB group was 0.67 (95%CI: 0.37 to 1.20; P = 0.176). Among 271 severe cases without IKF, the odds ratio of case-fatality rate was 0.42 (95CI%: 0.18 to 0.99; P = 0.046) for patients in the CCB group after adjustment of age, sex, and underlying diseases.

**Conclusions:** Hypertension is not just a chronic underlying comorbidity, but also a risk factor affecting the severity of COVID-19 and does play a critical role in worsening patients’ clinical outcomes. Therefore, hypertension management in patients with COVID-19 should be regarded as a major challenge in the diagnostic and therapeutic strategies.

**Trial registration:** N.A.

## Introduction

The infection of COVID-19 has swiftly spread worldwide. The clinical spectrum of COVID-19 patients appears to be wide, ranging from asymptomatic infection to mild, and to critical illness.^[1, 2]^ A significant portion of COVID-19 patients were reported to have at least one underlying complication when admitted to hospital, with hypertension, diabetes mellitus, and other cardiovascular diseases being the most common.^[3]^

There were about 20% of COVID-19 patients abruptly progressing from mild or normal condition to severe or critical illness, while the critical determinants remain to be identified. For early detection, early prediction, and early treatment, identifying the factors and biological indicators, in particular, those of early warning value for the development of disease progression and prognosis, are critical for clinical decision making and medical resource allocation.

In this article, we conducted an in-depth retrospective analysis of COVID-19 inpatients who were received and treated intensively in the early stage of the pandemic. Our aim was to reveal the factors of great influence on and predictive value of COVID-19 progression through the medical history, laboratory indicators and prognosis outcomes.

## Methods

### Study design and participants

This retrospective cohort study included 1031 COVID-19 inpatients from Tongji Hospital, Tongji Medical College of HUST (Wuhan, China). All patients were diagnosed with COVID-19 according to *Prevention and control Scheme for Novel Coronavirus Pneumonia* (5^th^ edition) published by National Health Commission of the People’s Republic of China^[4]^ and WHO interim guidance. A confirmed case of COVID-19 was defined as a positive result of serum antibody or real-time reverse-transcriptase–polymerase-chain-reaction (RT-PCR) assay of nasal or pharyngeal swab specimens. The primary outcome was the occurrence of cases with composite endpoints. The case-critically ill rate, the rate of case-fatality rate, and length of stay were included in the secondary outcomes.

The admission date of these hospitalized patients was from January 27, 2020, to March 8, 2020. The date cutoff for the study was March 30, 2020. The study was designed by the investigators and conducted in accordance with the principles of the Declaration of Helsinki. The Institutional Review Board of Tongji Hospital, Wuhan, China, approved this retrospective study and written informed consent was waived (No. TJ-C20200140). Data was analyzed and interpreted by the authors.

### Data collection

We obtained raw data regarding 1031 hospitalized COVID-19 patients in Tongji Hospital. Demographic data, medical history, clinical symptoms and signs, laboratory findings, chest computed tomography (CT), treatment and clinical outcomes were extracted from electronic medical records with standardized data collection forms for hospitalized COVID-19 patients. All data were doubled checked by two experienced physicians to ensure the accuracy of data collection. Disagreement between these two major reviewers was adjudicated by a third reviewer.

Demographic data included age and sex; clinical symptoms and signs included fever, cough, dyspnea, myalgia, diarrhea, chest congestion, heart rate, blood pressure and OOS; Laboratory findings from laboratory information system consist of a complete blood count, LDH, PT, APTT, Scr, eGFR, ALT, AST, serum albumin, D-dimer, NT-proBNP, TNI, CRP, ESR, IL-6, SARS-CoV-2 IgG, SARS-CoV-2 IgM and SARS-CoV-2 nucleic acid detection. Chest CT report data was obtained from image acquisition and communication systems.

Smoking history and baseline comorbidities (hypertension, DM, CHD, COPD, IKF, stroke and cancer) were extracted from medical history. Drug use condition (anti-hypertension, anti-coronary drugs, antiviral drugs, antibiotics, corticosteroids, gamma globulin, traditional Chinese medicine) and mechanical interventions (non-invasive ventilator, tracheal intubation, ECMO, CRRT) of COVID-19 patients during hospitalization were collected from medical advice. Information about the time from symptom onset to admission to hospital, severity of illness (normal/severe), and clinical outcomes (discharged or deceased) were obtained from the electronic medical system.

### Patient and Public Involvement statement

Patients and the public were not involved in the research.

### Study definitions

According to *Diagnosis & Treatment Scheme for Novel Coronavirus Pneumonia* (Trial 7^th^ edition) published by National Health Commission of the People’s Republic of China,^[5]^ the severity of COVID-19 inpatients was classified into mild, common, severe and critically ill. Acute cardiac injury (ACI) was diagnosed if serum levels of TNI were more than 0.342 μg/L for males and more than 0.156 μg/L for females. Impaired kidney function (IKF) was diagnosed if eGFR was less than 60 ml/min in the admission examination. The composite endpoint was the occurrence of ACI, IKF, or use of tracheal intubation. Different groups were divided on the basis of collected medical history. Patients with hypertension were included in the hypertension group, and patients without hypertension were included in the non-hypertension group. The simple hypertension group referred to patients with hypertension while without diabetes mellitus. The simple diabetes mellitus group referred to patients with diabetes mellitus while without hypertension. The hypertension and diabetes mellitus group referred to patients with both hypertension and diabetes mellitus. The others group referred to patients without hypertension and diabetes mellitus. Patients without hypertension, DM, CHD, COPD and cancer were included in the non-underlying disease group. ^[6]^ The CCB treatment mainly refers to the use of dihydropyridine long-acting calcium channel blockers during hospitalization.

### Statistical analysis

Continuous variables were presented as medians with standard deviation (SD) and compared with independent sample T test. Categorical variables were summarized as counts and percentages n (%) and compared with Pearson χ^2^ test. The missing data was removed. We used univariable and multivariable logistic regression models to look for insights into risk factors associated with incidence of critically ill cases, fatality cases and composite endpoints. A two-sided α of less than 0.05 was considered statistically significant. All the analyses were performed using SPSS (version 21.0) and GraphPad Prism (version 8.4.0) software.

## Results

There were 514 patients with chronic comorbidities in the total 1031 COVID-19 inpatients, accounting for 50%, two thirds of whom had one chronic comorbidities. The CFR (23.5%) of patients with chronic comorbidities was 2.8 times as that of patients with no underlying disease (8.5%). No difference was observed in CFR among patients with different number of chronic comorbidities.

There were 866 discharged patients and 165 deceased patients in the total 1031 COVID-19 inpatients. The clinical characteristics and prognosis conditions, including demographic data, clinical symptoms and signs, chronic comorbidities, and drug therapy, of these two groups are listed in Table 1. Among deceased patients, 73% had chronic comorbidities (Table 1), 56% (92/165) had hypertension or diabetes mellitus (Appendix Table 1), 47% had hypertension (Table 1).

**Table 1.**
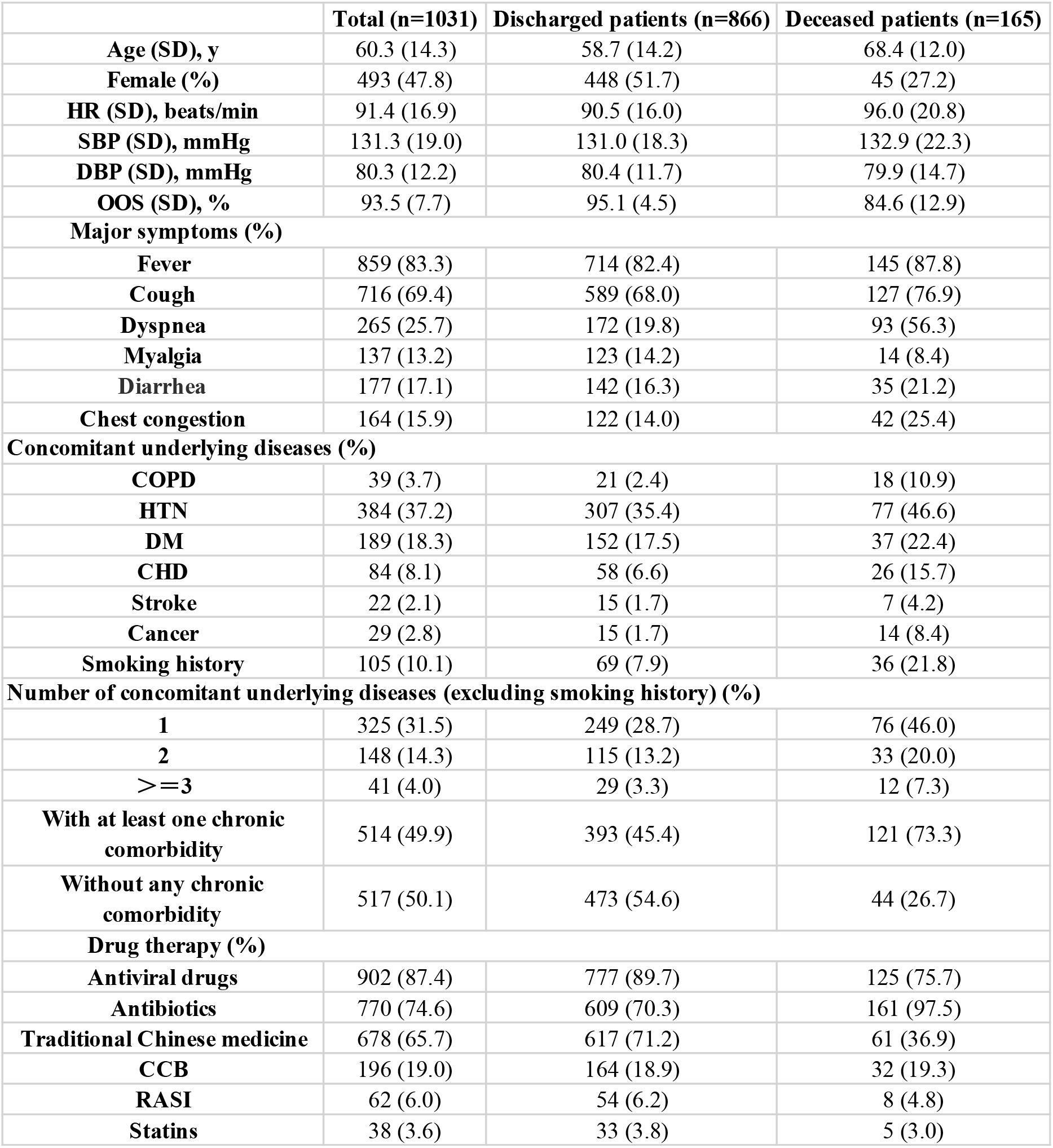
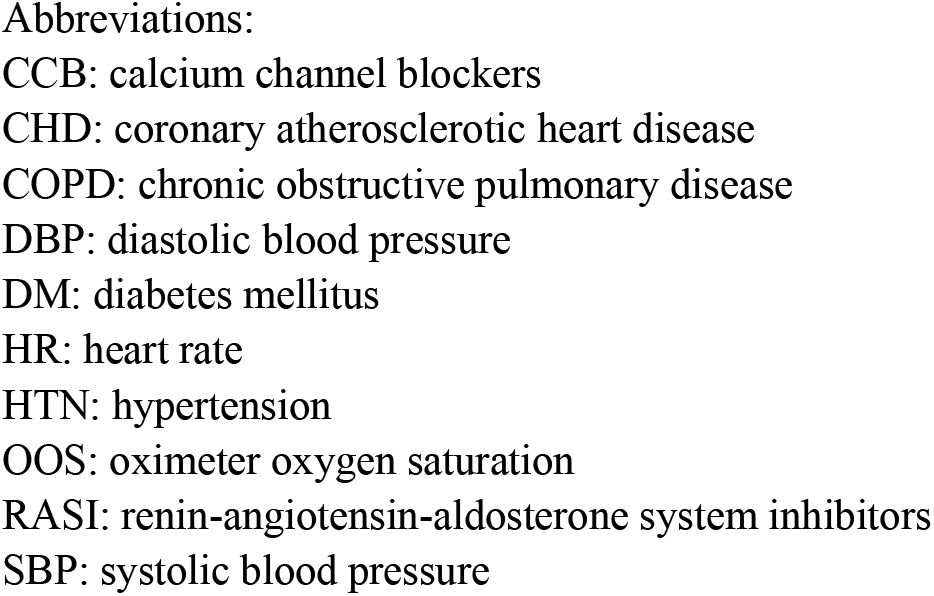
Comparison of clinical characteristics and prognosis conditions between discharged and deceased patients.

|  | **Total (n=1031)** | **Discharged patients (n=866)** | **Deceased patients (n=165)** |
| --- | --- | --- | --- |
| **Age (SD), y** | 60.3 (14.3) | 58.7 (14.2) | 68.4 (12.0) |
| **Female (%)** | 493 (47.8) | 448 (51.7) | 45 (27.2) |
| **HR (SD), beats/min** | 91.4 (16.9) | 90.5 (16.0) | 96.0 (20.8) |
| **SBP (SD), mmHg** | 131.3 (19.0) | 131.0 (18.3) | 132.9 (22.3) |
| **DBP (SD), mmHg** | 80.3 (12.2) | 80.4 (11.7) | 79.9 (14.7) |
| **OOS (SD), %** | 93.5 (7.7) | 95.1 (4.5) | 84.6 (12.9) |
| **Major symptoms (%)** |  |  |  |
| **Fever** | 859 (83.3) | 714 (82.4) | 145 (87.8) |
| **Cough** | 716 (69.4) | 589 (68.0) | 127 (76.9) |
| **Dyspnea** | 265 (25.7) | 172 (19.8) | 93 (56.3) |
| **Myalgia** | 137 (13.2) | 123 (14.2) | 14 (8.4) |
| **Diarrhea** | 177 (17.1) | 142 (16.3) | 35 (21.2) |
| **Chest congestion** | 164 (15.9) | 122 (14.0) | 42 (25.4) |
| **Concomitant underlying diseases (%)** |  |  |  |
| **COPD** | 39 (3.7) | 21 (2.4) | 18 (10.9) |
| **HTN** | 384 (37.2) | 307 (35.4) | 77 (46.6) |
| **DM** | 189 (18.3) | 152 (17.5) | 37 (22.4) |
| **CHD** | 84 (8.1) | 58 (6.6) | 26 (15.7) |
| **Stroke** | 22 (2.1) | 15 (1.7) | 7 (4.2) |
| **Cancer** | 29 (2.8) | 15 (1.7) | 14 (8.4) |
| **Smoking history** | 105 (10.1) | 69 (7.9) | 36 (21.8) |
| **Number of concomitant underlying diseases (excluding smoking history) (%)** |  |  |  |
| **1** | 325 (31.5) | 249 (28.7) | 76 (46.0) |
| **2** | 148 (14.3) | 115 (13.2) | 33 (20.0) |
| **>=3** | 41 (4.0) | 29 (3.3) | 12 (7.3) |
| **With at least one chronic comorbidity** | 514 (49.9) | 393 (45.4) | 121 (73.3) |
| **Without any chronic comorbidity** | 517 (50.1) | 473 (54.6) | 44 (26.7) |
| **Drug therapy (%)** |  |  |  |
| **Antiviral drugs** | 902 (87.4) | 777 (89.7) | 125 (75.7) |
| **Antibiotics** | 770 (74.6) | 609 (70.3) | 161 (97.5) |
| **Traditional Chinese medicine** | 678 (65.7) | 617 (71.2) | 61 (36.9) |
| **CCB** | 196 (19.0) | 164 (18.9) | 32 (19.3) |
| **RASI** | 62 (6.0) | 54 (6.2) | 8 (4.8) |
| **Statins** | 38 (3.6) | 33 (3.8) | 5 (3.0) |

Abbreviations:
CCB: calcium channel blockers
CHD: coronary atherosclerotic heart disease
COPD: chronic obstructive pulmonary disease
DBP: diastolic blood pressure
DM: diabetes mellitus
HR: heart rate
HTN: hypertension
OOS: oximeter oxygen saturation
RASI: renin-angiotensin-aldosterone system inhibitors
SBP: systolic blood pressure

According to Table 1, logistic regression analyses were conducted with factors like the elderly (patients >= 65 years old), males, and a variety of underlying diseases included (Appendix Table 2). All the underlying diseases were revealed to show relations to case fatality except for diabetes mellitus (P > 0.05). The multivariable logistic regression analysis still showed the relations between COPD, CHD, stroke, cancer, smoking history and case fatality after adjustment. The corresponding odds ratio of CFR were 2.69 (95%CI: 1.32 to 5.49; P=0.006), 2.04 (95%CI: 1.18 to 3.55; P=0.011), 2.70 (95%CI: 1.00 to 7.31; P=0.050), 8.01 (95%CI: 3.55 to 18.07; P<0.001), 2.05 (95%CI: 1.25 to 3.38; P=0.005) respectively, while the odds ratio for hypertensive patients was 1.05 (95%CI: 0.72 to 1.54; P>0.05). The adjusted odds ratio of CFR were 2.94 (95%CI: 2.00 to 4.33; P<0.001) and 2.47 (95%CI: 1.65 to 3.70; P<0.001) respectively for elderly patients and male patients.

### A. The subgroup analysis of prognosis in patients with underlying hypertension and diabetes mellitus

There were 573 patients included in the others group (Others), 269 in the simple hypertension group (HTN), 74 in the simple diabetes mellitus group (DM) and 115 in the hypertension and diabetes mellitus group (HTN+DM). Multiple indicators including CCIR, CFR, the incidence of ACI, IKF, and tracheal intubation, the use of non-invasive ventilator, ECMO and CRRT, were compared among these four groups (Figure 1). The CCIR and CFR were relatively higher in groups with chronic comorbidities. Compared to Others group, various prognostic indicators were significantly higher in HTN group and DM group. Patients in HTN+DM group showed higher incidence of ACI, IKF and tracheal intubation and the use of non-invasive ventilator.

**Figure 1.**
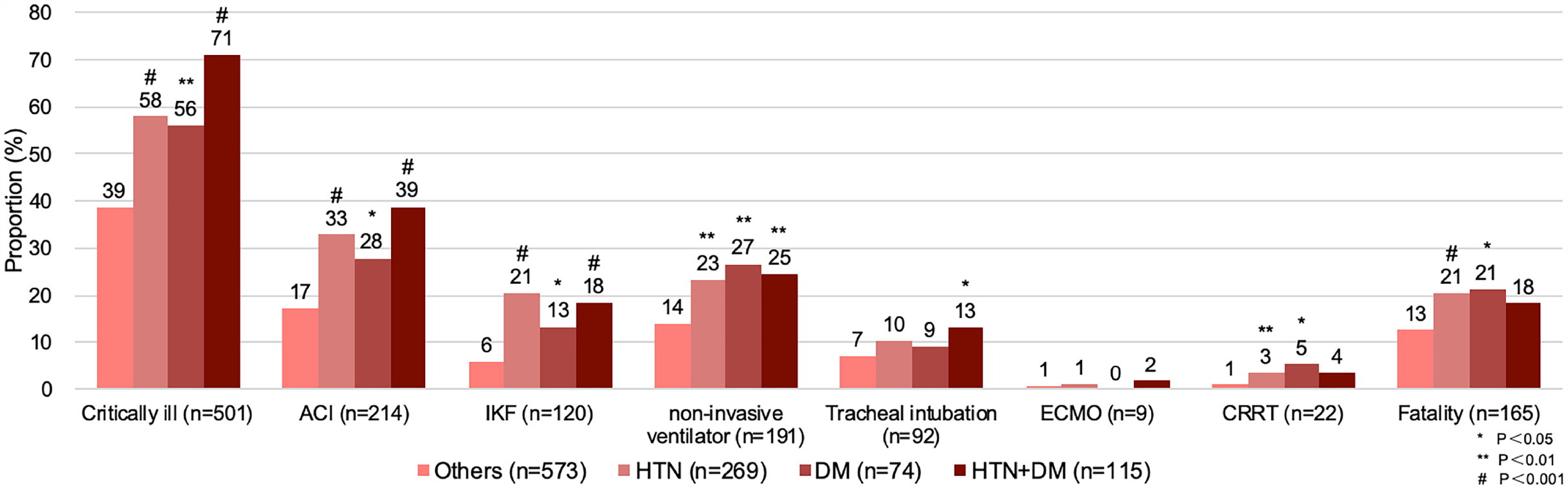
Comparison of various clinical outcomes among groups with and without chronic comorbidities. Abbreviations: ACI: acute cardiac injury CRRT: continuous renal replacement therapy DM: diabetes mellitus ECMO: extracorporeal membrane oxygenation HTN: hypertension IKF: impaired kidney function

Odds ratio of three major prognostic indicators (incidence of severe cases, fatality cases and cases with composite endpoints) were adjusted by age (Figure 2) in COVID-19 inpatients with HTN, DM and HTN+DM group, and showed that patients with simple hypertension had higher risk than those with simple diabetes mellitus early in the disease. The Odds ratio of occurrence of composite endpoints was 1.53 (95%CI: 1.07 to 2.17; P=0.019) in the HTN group and 1.55 (P<0.001) in the HTN+DM group (Figure 2, Appendix Table 1). Odds ratio of case fatality was 1.18 (95%CI: 0.99 to 1.41; P=0.063) in the HTN+DM group, indicating its increasing trend in death. (Figure 2).

**Figure 2.**
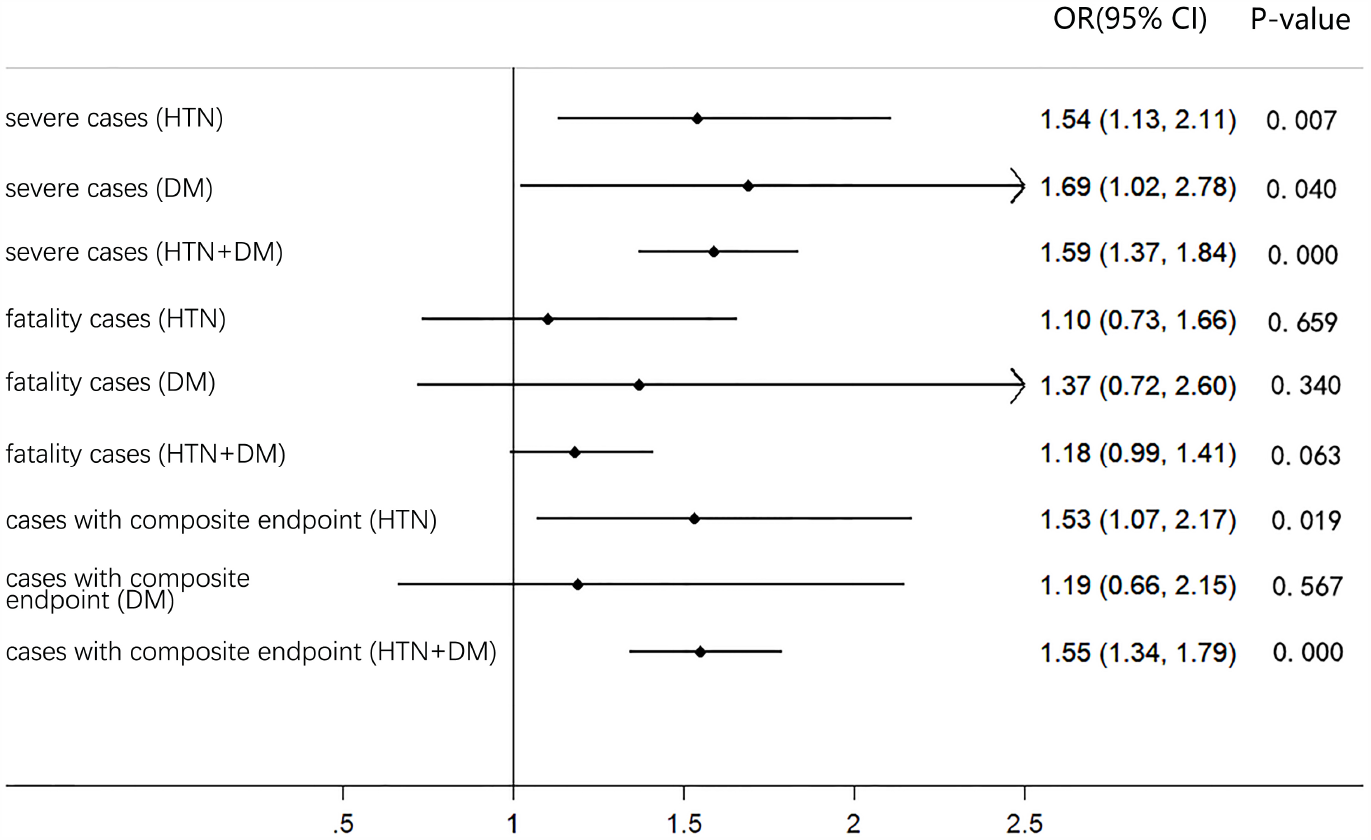
Comparison of age-adjusted odds ratio of three major prognostic indicators for the HTN group, DM group, and HTN+DM group (compared to Others group) Abbreviations: DM: diabetes mellitus HTN: hypertension

### B. The subgroup analysis of case fatality in patients with underlying hypertension

The total of 1031 COVID-19 inpatients were divided into four groups, ie the hypertensive discharged group (307 patients), hypertensive deceased group (77 patients), non-hypertensive discharged group (559 patients) and non-hypertensive deceased group (88 patients). Demographic data (age, sex), clinical signs (OOS, HR, SBP, DBP, MAP) and laboratory indicators (NEU, LYM, PLT, CRP, IL-6, LDH, D-dimer, eGFR) were compared among these four groups, as shown in Table 2. There was no significant difference observed between discharged and deceased patients in the hypertension and non-hypertension group in sex, OOS, HR, and many other laboratory indicators. However, the indicator of eGFR was significantly lower in the hypertension deceased group than the non-hypertension deceased group (66.9±25.7 vs. 78.3±25.9, P=0.005).

**Table 2.**
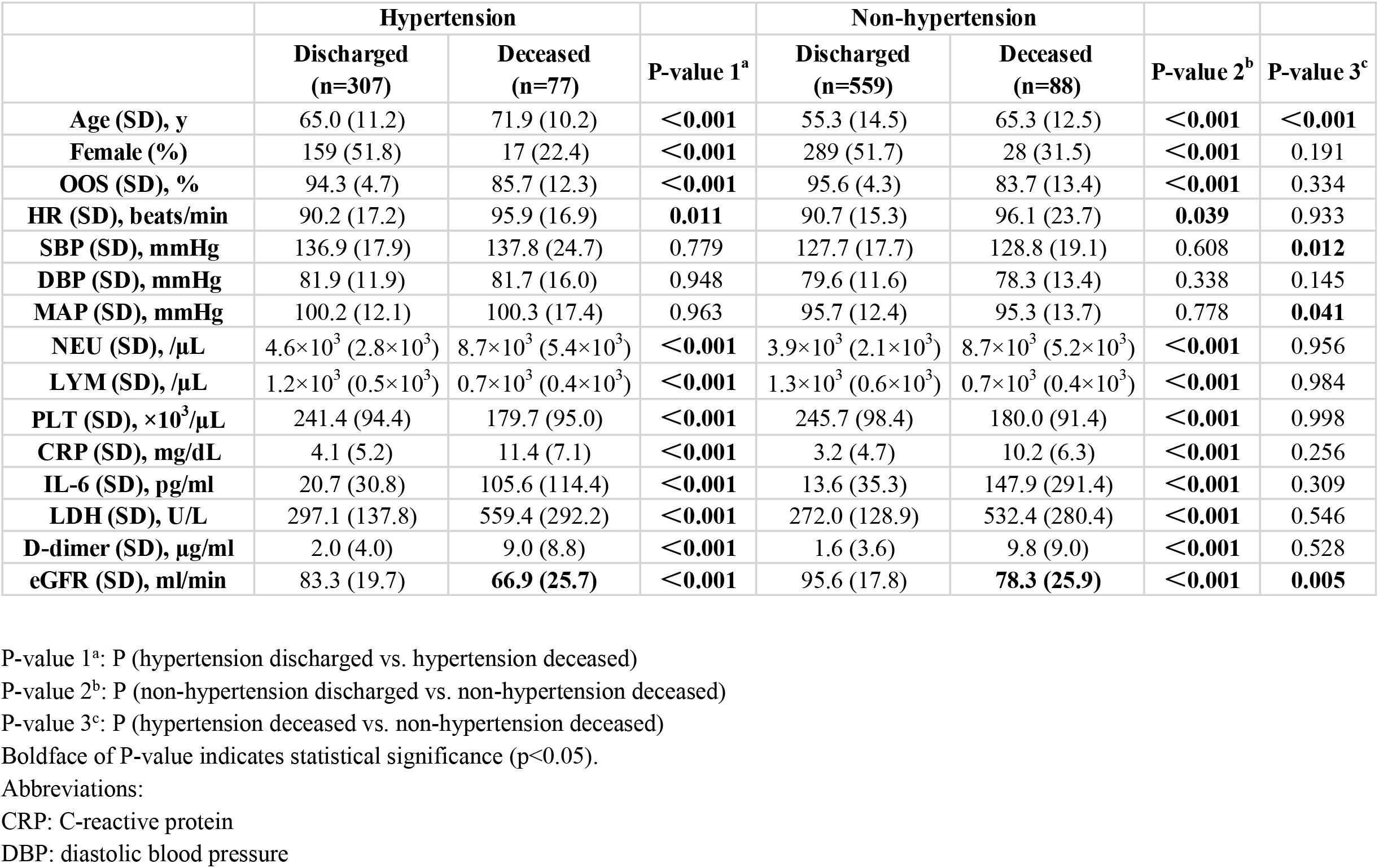
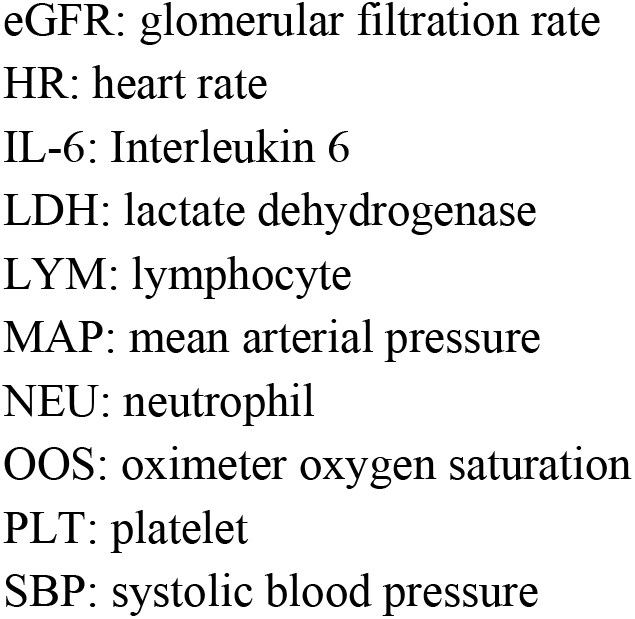
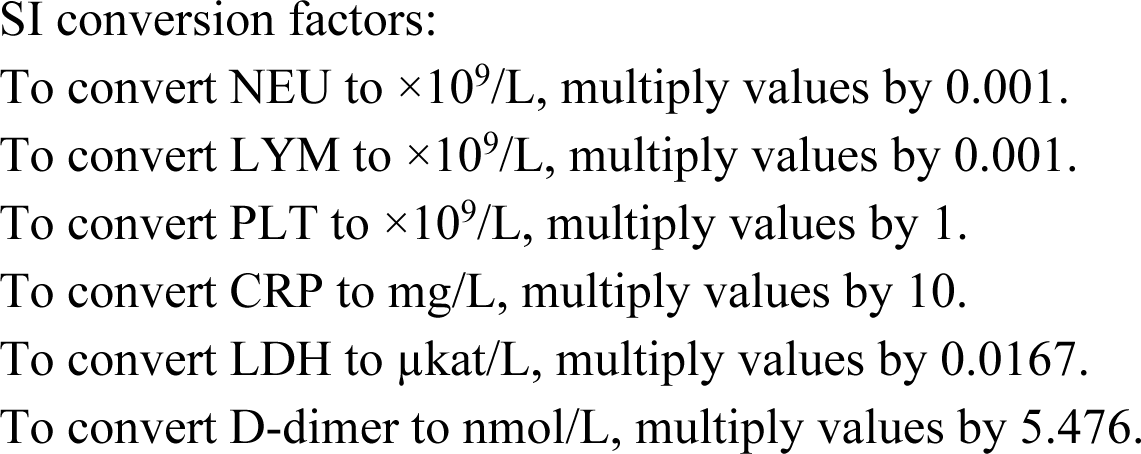
Comparison of illness condition of discharged and deceased patients in the hypertension and non-hypertension group.

### C. The subgroup analysis of senility in patients with underlying hypertension

There were 419 patients in the non-hypertensive non-senile group, 150 patients in the hypertensive non-senile patients, 228 patients in the non-hypertensive senile group, and 234 patients in the hypertensive senile group, when all patients with hypertension were divided according to age (the age of 65). Demographic data, clinical signs, laboratory indicators, and CCIR, CFR1, CFR2 were compared among the above four groups (Appendix Table 3). The results showed that the CCIR in the hypertensive senile group was significantly higher than the non-hypertensive senile group (71% vs. 51%, P<0.001). Different from non-senile patients, senile patients showed significantly difference only in the indicator of CRP, LDH, and eGFR between the hypertension group and non-hypertension group except for age and blood pressure. By further comparing the probability of survival among the four groups above (Figure 3), we found that the probability of survival in the hypertensive senile group continuously declined from the very start of hospitalization, lowest among four groups. The CFR of the hypertensive senile group was 26.9%.

**Figure 3.**
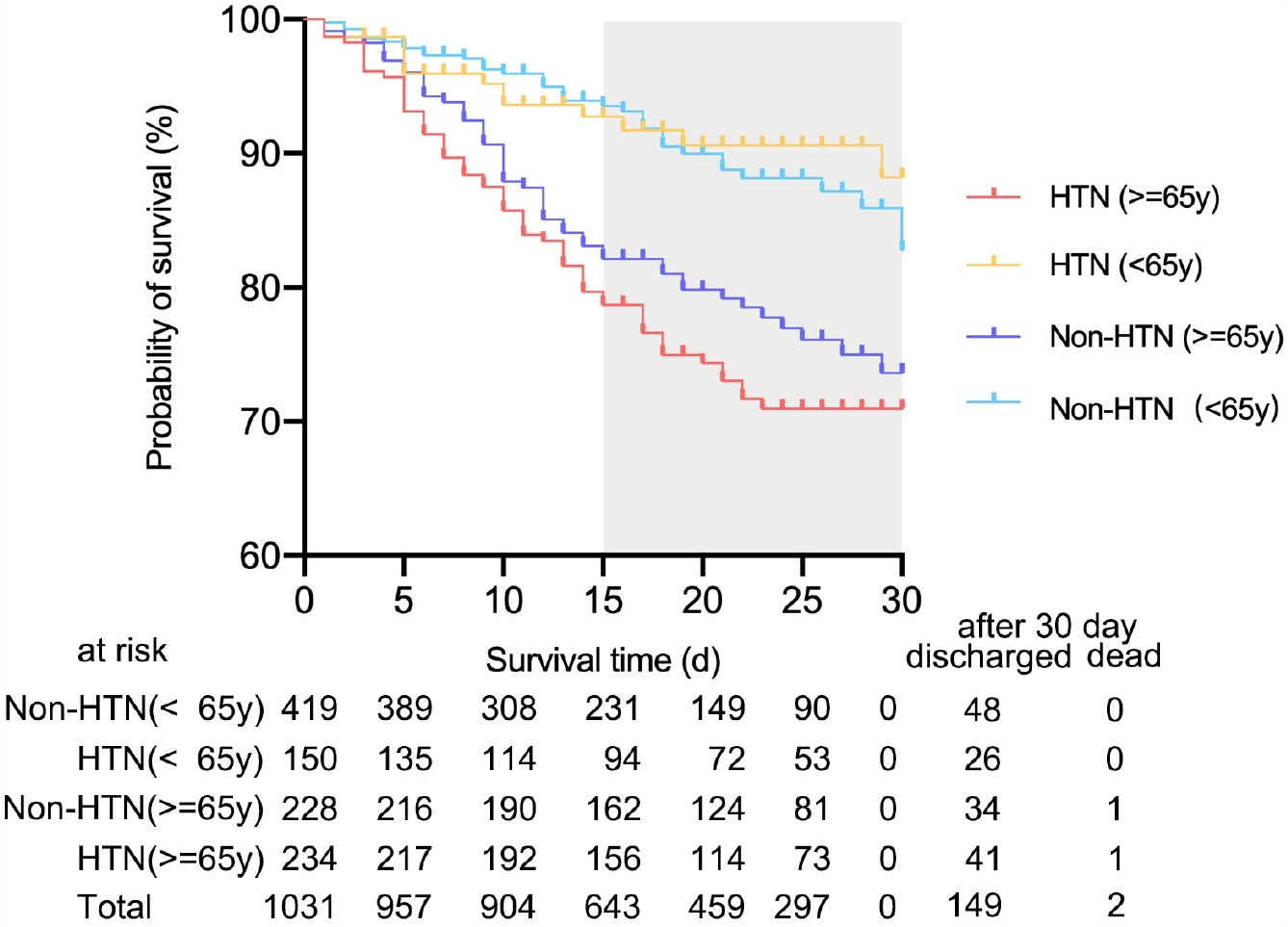
The probability of survival in 30 days among patients with different ages in the hypertension and non-hypertension group. Abbreviations: HTN: hypertension Non-HTN: non-hypertension

### D. The subgroup analysis of drug therapy for hospitalized patients

Among all 1031 hospitalized patients, 196 patients were treated with CCB during hospitalization, and 36 patients deceased (CFR=16%), the same as those not treated with CCB (133/835), while with longer LOS (23.7 vs. 20.8, P = 0.001). (Appendix Table 4) The longer LOS may be associated with more severe conditions before hospitalization in patients treated with CCB. The proportion of the elderly, patients with hypertension, and other underlying diseases were all higher in the CCB group compared to the non-CCB group. No significant difference was found in the proportion of males and the indicator of OOS between patients in these two groups. (Appendix Table 4)

Of 14 patients taking CCB during hospitalization in 328 low-risk cases, no fatality cases occurred (Figure 4), and there was a decreasing trend in LOS (13.2 vs. 17.9, P=0.066). (Appendix Table 4) CCB groups were observed to show longer LOS across high-risk groups of four different populations, suggesting that patients in these groups were at higher “risk of virus”. However, the CFR of CCB groups in all groupings was relatively lower. (Appendix Table 5)

**Figure 4.**
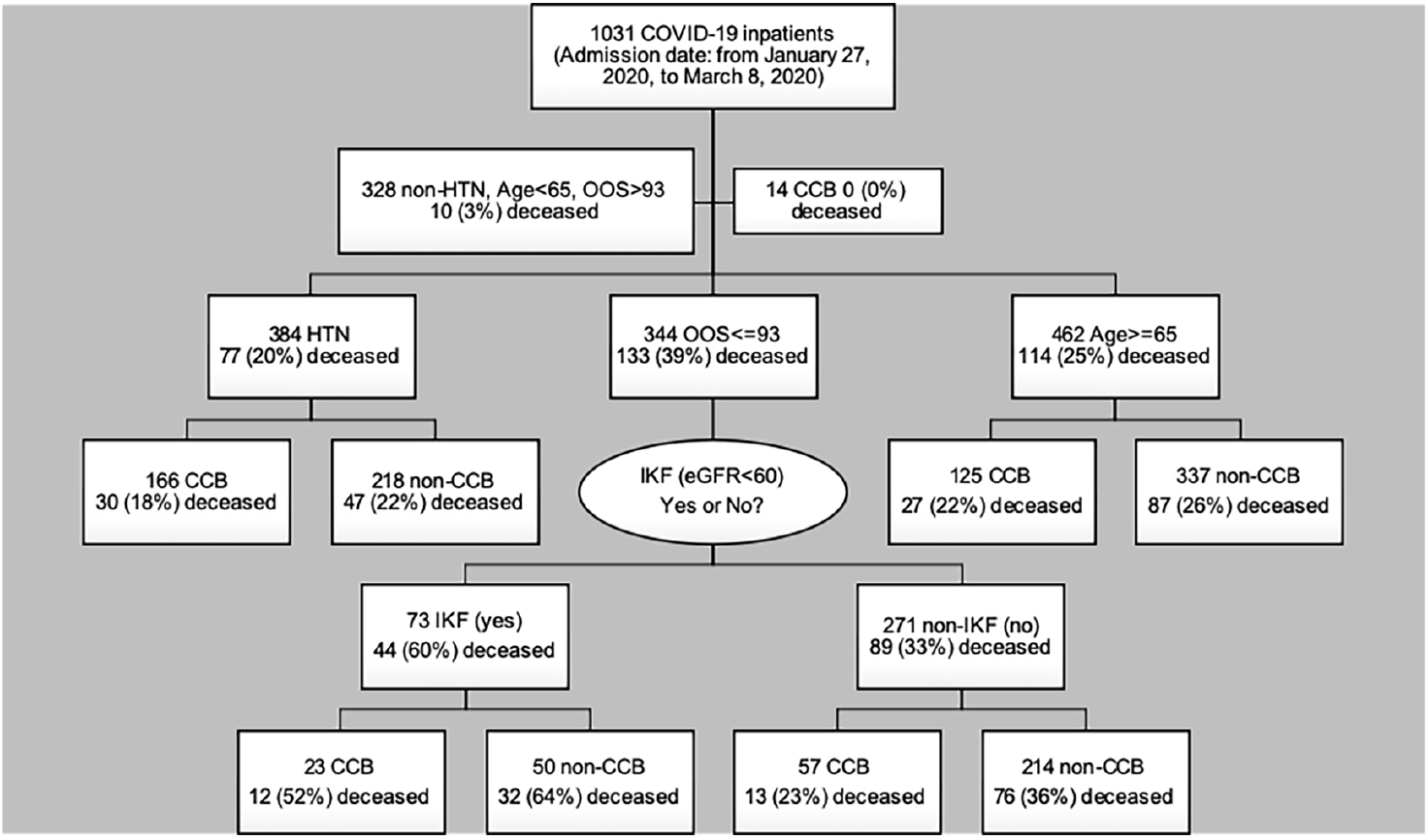
The subgroup analysis of different in-hospital populations in the CCB group and the non-CCB group. Abbreviations: CCB: calcium channel blockers HTN: hypertension IKF: impaired kidney function Non-HTN: non-hypertension OOS: oximeter oxygen saturation

Among 384 patients with hypertension, 30 of 166 receiving in-hospital treatment of CCB deceased, with CFR lower than that of the non-CCB group (18% vs. 22%). (Figure 4) By comparing various clinical characteristics between the CCB group and the non-CCB group (Appendix Table 6), only age, the proportion of patients combining with DM, IKF, and receiving RASI treatment during hospitalization were observed to be significantly different. After adjustment for various relevant factors (Appendix Table 6), the odds ratio of CFR for hypertensive patients in the CCB group was 0.67 (95%CI: 0.37 to 1.20, P = 0.176). 57 were treated with CCB during hospitalization and 13 deceased among 271 severe cases without IKF, with CFR lower than that of the non-CCB group (23% vs. 36%). (Figure 4) The odds ratio of CFR for severe cases without IKF in the CCB group was 0.42 after adjustment for age, sex, and underlying diseases (95%CI: 0.18 to 0.99, P = 0.046), which was consistent with the result of an online study with small sample size. (Appendix Table 7)

## Discussion

The clinical manifestations of COVID-19 patients can range from asymptomatic infection to mild, and to critical illness requiring ventilators and even ECMO to sustain lives.^[1, 2]^ Wuhan scholars in China first reported that about half of the patients with severe infection were complicated with chronic comorbidities such as hypertension, diabetes mellitus or coronary heart diseases.^[7]^ Among those infected with underlying diseases, about 38% progressed to severe cases.^[7]^ About 60% of severe cases with chronic comorbidities deceased in hospital.^[7]^ However, whether chronic comorbidities can be regarded as a risk factor affecting the severity of COVID-19 remains controversial at that time.

Recently, there were many studies reporting that age and concomitant diseases might aggravate the disease progression and affect clinical outcomes among COVID-19 patients. COVID-19 patients with pre-existing complications were observed to show higher CFR-10.5%, 7.3%, 6.3%, 6.0%, 5.6% for patients with cardiovascular diseases, diabetes mellitus, chronic respiratory diseases, hypertension, and cancer respectively.^[6]^ It has been reported that the greater the number of underlying diseases is combined, the greater the severity of COVID-19.^[3]^ The retrospective analyses of COVID-19 patients in Zhejiang Province and in 31 province across mainland China revealed that underlying hypertension was an independent risk factor for severe cases, even after adjustment for age.^[3, 8]^ Among 1043 COVID-19 patients admitted to ICU with available data in Italy, 49% had underlying hypertension. After excluding patients still in ICU as of the deadline of the study, the CFR for patients with and without hypertension was 70% and 47%.^[9]^ The result of the population-based cohort study in England supported that patients with hypertension should be included in the high-risk group.^[10]^

In our retrospective study, it was found that the determinants of the severity of COVID-19 were closely related to the age, male, smoking history, and chronic comorbidities. Chronic comorbidities, such as hypertension and diabetes mellitus, are major risk factors affecting disease progression and prognosis, though they were revealed to show no relations with CFR after adjustment for multiple factors. Patients with simple hypertension may be at higher risk than those with simple diabetes mellitus at early stages of the disease after adjustment for age, and senile hypertensive patients are at high risk for early progression and prognosis of COVID-19. The CFR of senile hypertensive patients was 26.9%.

Why do chronic comorbidities, such as hypertension, increase the risk of the disease progression and its prognosis? It was very important and should be investigated. Now, an article named *Viral and host factors related to the clinical outcome of COVID-19* was published online,^[11]^ to analyze viral genome variation from sample collection of 326 patients in Shanghai Public Health Center. No significant variation of viral sequences was observed in severe cases of COVID-19, therefore ruling out the possibility of mutation of SARS-CoV2 itself. Multiple major laboratory indicators were compared among discharged and deceased patients with and without hypertension, there was no significant difference observed in general. There was also no significant difference found in SBP and MAP among these four groups. The results suggest that it is likely due to the pathophysiological status and target organ damage in patients with hypertension, such as RAS system and the laboratory indicator of eGFR, rather than the blood pressure itself that makes hypertension as a risk factor of earlier illness development and prognosis. A single-center study including 417 hospitalized cases in in Kuwait published online has found that their dynamic profiling of eGFR in COVID-19 ICU patients highlights potential role of renal markers in forecasting disease outcome and may identify patients at risk of poor outcome, which is consistent with the results of our study.^[12]^

It is now known that the infection of SARS-CoV2 needs its combination with ACE2 and TMPRSS2 of human alveolar epithelial cells type 2 or other tissues and subsequent replication and multiplication. The amount of virus leads to increased expression of ACE2 in neighboring cells, and then affects macrophages with inflammatory cytokines released.^[13]^ Consequently, the increased expression and activity of ACE2 in tissue could be identified as the critical factor of patients susceptibility and disease progression. The Huanggang study, published online, conducted serological tests of ACE2 in 20 COVID-19 patients and 20 non-COVID-19 patients, revealing that the serum level of ACE2 increased 12 hours after infection and remained high 48 hours after infection.^[14]^ The expression of ACE2 is only mildly expressed in lung tissue of healthy human, however, Brazilian researchers recently found that the expression of ACE2 in lung tissue was up-regulated in COVID-19 patients with underlying diseases,^[15]^ including hypertension, which might provide a reasonable explanation for the increased severity in some COVID-19 patients with underlying complications.

To our knowledge, this study is a retrospective report mainly focusing on patients with hypertension, which accounts for the highest proportion of COVID-19 patients at risk. As early targeted interventions will prevent the disease from further progressing, a reasonable management strategy of hypertension is of great necessity, and might help to improve clinical outcomes in hospitalized COVID-19 patients at high risk. In a previously published retrospective cohort study in Tongji Hospital, Tongji Medical College of HUST (Wuhan, China) focusing on patients admitted after January 14, 2020, ^[16]^ there were 75 deceased cases of 306 patients with baseline hypertension (CFR=24.5%). Meanwhile, there was another study conduted in New York City with 384 of 1366 hypertensive patients deceased until April 4, 2020 (CFR=28.1%). ^[17]^However, 77 deceased cases occurred among 384 inpatients with hypertension in our retrospective study with CFR being 20.1%, lower than that of two above studies. The difference might be associated with increased CCB in-hospital treatment according to expert recommendations announced by Hubei Cardiovascular Internal Medicine Medical Quality Control Center and Wuhan Medical Association Cardiovascular Disease Credit Association on February 1, 2020. ^[18]^ Subgroup analyses of different populations in our study suggested that the main benefit of CCB in-hospital treatment might come from patients in severe conditions on admission. Among 344 severe hospitalized cases with OOS less than 93% (including 93%), 25 of 80 in the CCB group deceased (CFR=31%) and 108 of 264 deceased in the non-CCB group (CFR=41%), which was consistent with the result of an online study with small sample size. The study indicated a beneficial effect of amlodipine besylate in reducing the CFR of hypertensive patients, with the CFR being 6.8% (3/44) in amlodipine besylate treated group compared to 26.1% (12/46) in non-amlodipine besylate treated group to 6.8% (3/44) (P = 0.022). ^[19]^

Our retrospective cohort study has some limitations. Firstly, only admission examination for the indicator of eGFR was captured in the collected data, and the overall level of IKF might therefore be underestimated. Secondly, the information of patients prior to admission was not put into the medical history, so it could not be determined whether the patients continued to take or changed the medication during hospitalization. Thirdly, there was no national unified standard for the normal reference values of some laboratory indicators, thus we followed the standards set by the hospital. Finally, the data entry of underlying diseases was based on the inquiries of patients enrolled in the hospital and might be underestimated due to the influence of awareness rate.

## Data Availability

The raw data was extracted from electronic medical records with standardized data collection forms in Tongji Hospital (Wuhan, China), which had been approved by the Research Ethics Commission of Tongji Hospital, Tongji medical college of HUST. The data used to support the findings of this study are available from the corresponding author upon request.

## Nonstandard Abbreviations and Acronyms

ACI: Acute cardiac injury
ALT: Alanine aminotransferase
APTT: Activated partial thromboplastin time
AST: Aspartate aminotransferase
CCB: Calcium channel blockers
CCIR: Case-critically ill rate
CDC: Chinese Center for Disease Control and Prevention
CFR: Case-fatality rate
CHD: Coronary atherosclerotic heart disease
COPD: Chronic obstructive pulmonary disease
COVID-19: Coronavirus disease 2019
CRPC: -reactive protein
CRRT: Continuous renal replacement therapy
DM: Diabetes mellitus
ECMO: Extracorporeal membrane oxygenation
eGFR: Glomerular filtration rate
ESR: Erythrocyte sedimentation rate
IKF: Impaired kidney function
IL-6: Interleukin 6
LDH: Lactate dehydrogenase
LOS: Length of stay
MAP: Mean arterial pressure
NT-proBNP: N-terminal pro-brain natriuretic peptide
OOS: Oximeter oxygen saturation
PT: Prothrombin time
RAS: Renin-angiotensin-aldosterone system
RASI: Renin-angiotensin-aldosterone system inhibitors
SARS-CoV-2 IgG: SARS-CoV-2 Immunoglobulin G
SARS-CoV-2 IgM: SARS-CoV-2 Immunoglobulin M
Scr: Serum creatinine
TNI: Troponin I
WHO: World Health Organization

## Authors’ Contributors

Hesong Zeng, Weizhong Zhang, and Yin shen conceived the study and its design, had full access to all the data in the study, and take responsibility for the integrity of the data and accuracy of the data analysis. Xingwei He, Yuxin Du, and Yan Tong organised and entered data. Tianlu Zhang, Yuxin Du, and Xueli Wang contributed to data analyses. Hesong Zeng, Weizhong Zhang, Yin Shen, Xingwei He, and Tianlu Zhang participated in discussion and made crucial suggestions for interpreting the findings. Weizhong Zhang and Tianlu Zhang drafted the manuscript.

## Transparency declaration & Independence statement

The correspongding author, Yin Shen, affirms that the manuscript is an honest, accurate, and transparent account of the study being reported and that no important aspects of the study have been omitted. The independence of researchers from study sponsor was confirmed

## Acknowlegements

We thank all COVID-19 patients for their participation, our colleagues for their assistance, and the funding source for their involvement and support.

